# Body dissatisfaction widens the racial disparities of Benzophenone-3, a chemical biomarker of personal care and consumer product usage

**DOI:** 10.1101/2024.08.26.24312258

**Authors:** Vy Kim Nguyen, Samuel Zimmerman, Justin Colacino, Olivier Jolliet, Chirag J Patel

## Abstract

**Background:** Body dissatisfaction can drive individuals to use personal care products, exposing themselves to Benzophenone-3 (BP3). Yet, no study has examined the link between body dissatisfaction and elevated chemical exposures.

**Objectives:** Our study examines how body dissatisfaction impacts the racial differences in BP3 exposures.

**Methods:** Using NHANES 2003-2016 data for 3,072 women, we ascertained body dissatisfaction with a questionnaire on weight perception. We ran two generalized linear models with log10-transformed urinary concentrations of BP3 as the outcome variable and the following main predictors: one with race/ethnicity and another combining race/ethnicity and body dissatisfaction. We also conducted stratified analyses by race/ethnicity. We adjusted for poverty income ratio, BMI, urinary creatinine, and sunscreen usage.

**Results:** BP3 levels in Mexican American, Other Hispanic, Other Race, non-Hispanic White, and non-Hispanic Asian women were on average 59%, 56%, 33%, 16%, and 9% higher, respectively, compared to non-Hispanic Black women. Racial differences in BP3 levels are accentuated with body dissatisfaction. For example, Other Hispanic women perceiving themselves as overweight had 69% higher BP3 levels than non-Hispanic Black women (p-value = 0.01), while those perceiving themselves as at the right weight had 32% higher levels (p-value = 0.31). Moreover, minority women perceiving themselves as overweight tended to have higher BP3 levels than those who do not. For example, BP3 levels in Other Hispanic women perceiving themselves as overweight are significantly higher compared to those who do not (73%, p-value = 0.03). In contrast, such differences in the non-Hispanic White women are minimal (-0.5%, p-value = 0.98).

**Discussion:** Minority women with body dissatisfaction show elevated BP3 exposure independent of sunscreen usage, implying that their elevated exposures may stem from using other personal care and consumer products. Further research is needed to determine if increases of exposure to potential toxicants occur among minority women with body dissatisfaction.

**Highlights:**

⍰ First integration of measures of body dissatisfaction with national chemical biomonitoring data
⍰ Analyzed data from a diverse US nationally representative sample of 3,072 women
⍰ Minority women with body dissatisfaction show higher BP3 levels independent of sunscreen use and BMI
⍰ Minimal differences in BP3 levels by body dissatisfaction in non-Hispanic White women
⍰ Developed a visualization tool to show how racial disparities widen due to body dissatisfaction

## 1. Introduction

According to the Dove Self-Esteem project, body dissatisfaction, caused by toxic beauty standards, is estimated to be a $305 billion dollar public health crisis^1^. Body dissatisfaction is the negative attitude towards one’s own body because of a perceived difference between one’s appearance^2,3^ and the ideal body image promoted by society^4^. Such standards emphasize features like thinness, straight hair, and lighter skin. Body dissatisfaction approximately affects 45 million Americans or 16% of the US population aged 10 years and older^1,5^. Women and girls shoulder 63% of the total cost, amounting to $177 billion^1^. While many different health outcomes were considered (e.g., mental health issues, substance abuse), the report does not account for the impact of being exposed to toxicants through the use of cosmetic products to conform to these standards^1^. This oversight likely results in underestimating the true cost by overlooking the toxic effects of chemical exposures.

The environmental injustice of beauty is the internalization of racialized beauty standards, which may drive women of color to use cosmetic products and consequently be exposed to toxicants with adverse health outcomes^6^. These standards often lead to body image dissatisfaction, prompting individuals to use personal care products like chemical straighteners and skin lighteners to conform to societal norms. For instance, a Manhattan study on women of color revealed that the primary motivation behind using such cosmetic products is to attain a specific standard of beauty^7^. Additionally, dissatisfaction with body image, rooted in beauty standards, predicts more frequent makeup usage, increased time and money spent on makeup^8^, and appearance management behaviors^9^. Appearance management behaviors involve practices (e.g., using cosmetics) to maintain an ideal body image or enhance physical appearances. Using such products can expose individuals to environmental toxicants, implicated in serious health consequences such as skin reactions^10^, reproductive cancers^11,12^, and adverse birth outcomes^10^. Despite the risk, there is no published work on the link between body dissatisfaction and environmental toxicant exposures, and in turn, the extent of the chemical in the body. Additionally, the association between body dissatisfaction and chemical exposures has not been studied with respect to race/ethnicity. Understanding how body dissatisfaction is tied to elevated chemical exposures can help lead to intervention on building body satisfaction. By fostering body satisfaction, individuals may be less inclined to use cosmetic products, thus reducing their exposure to environmental toxicants and mitigating their adverse effects, particularly within vulnerable populations.

Due to its properties in absorbing and stabilizing ultraviolet light, Benzophenone-3 is commonly used in plastic products and as a sunscreen ingredient in personal care products to minimize damage and degradation from sunlight exposure. BP3 has been detected at concentrations that are 10 times higher compared to other chemical UV filters^13^. Furthermore, BP3 is detected in the urine of 96.95% of participants from National Health and Nutrition Examination Survey (NHANES), indicating widespread exposure to BP3 in the U.S. population^14^. The ubiquity of BP3 is compounded by its potential toxic effects^15^ related to skin^16,17^, kidney^18^, brain^19^, endocrine system^20^, hematological system^21^, reproductive system^22,23^, and neonatal development during pregnancy^24^. These associations necessitate the identification of populations most vulnerable to elevated BP3 exposures. The non-Hispanic White population has the highest biomarker levels of BP3, likely due to elevated sunscreen usage^14,25^. However, these studies did not consider how the racial disparities of BP3 would change when considering sunscreen usage. Studying racial differences in BP3 exposures while accounting sunscreen usage will help identify other BP3-containing products that contribute to these disparities.

Our goal is to examine how body dissatisfaction changes the racial disparities of exposures to BP3 after accounting for sunscreen usage. Our hypothesis is that minority women with body dissatisfaction tend to have higher biomarker levels of BP3 than those who do not have body dissatisfaction, even after adjusting for sunscreen usage. This would suggest that elevated BP3 levels in minority women with body dissatisfaction is from usage of other personal care and consumer products. To achieve our goal and test our hypothesis, we used data from NHANES 2003-2016. Our specific objectives are (1) characterizing the differences in biomarker levels of BP3 by race and body dissatisfaction, (2) evaluating how these differences change when accounting for sunscreen usage, and (3) quantifying how sunscreen usage is not the sole contributor of BP3 levels.

## 2. Methods

### 2.1 Study Population

CDC National Center for Health Statistics (NCHS) designed NHANES as a cross-sectional study to collect data on the demographics, dietary, chemical exposures, physiological indicators, and health-related questionnaires in the non-institutionalized, civilian US population. For this analysis, we used the Continuous NHANES data and started with our harmonized and curated version 1988-2018 (N = 135,310)^26^. After our curation process, we end with data from 2003-2016. We excluded participants who have missing data for weight perception (N = 71,947). As participants who perceived themselves as underweight comprised one-tenth of the other categories for weight perception (perceived at the right weight and perceived overweight), we excluded participants who perceived themselves as underweight (N = 3,913). We excluded male participants (N = 28,314). We excluded participants sequentially based on having missingness on the following: BP3 levels (N = 24,507), sunscreen usage (N = 3,299), poverty income ratio (PIR) (N = 231), body mass index (BMI) (N = 26), and urinary creatinine (N = 1). We have a final sample size of 3,072 female participants. These exclusion criteria are delineated in Figure 1.

**Figure 1.**
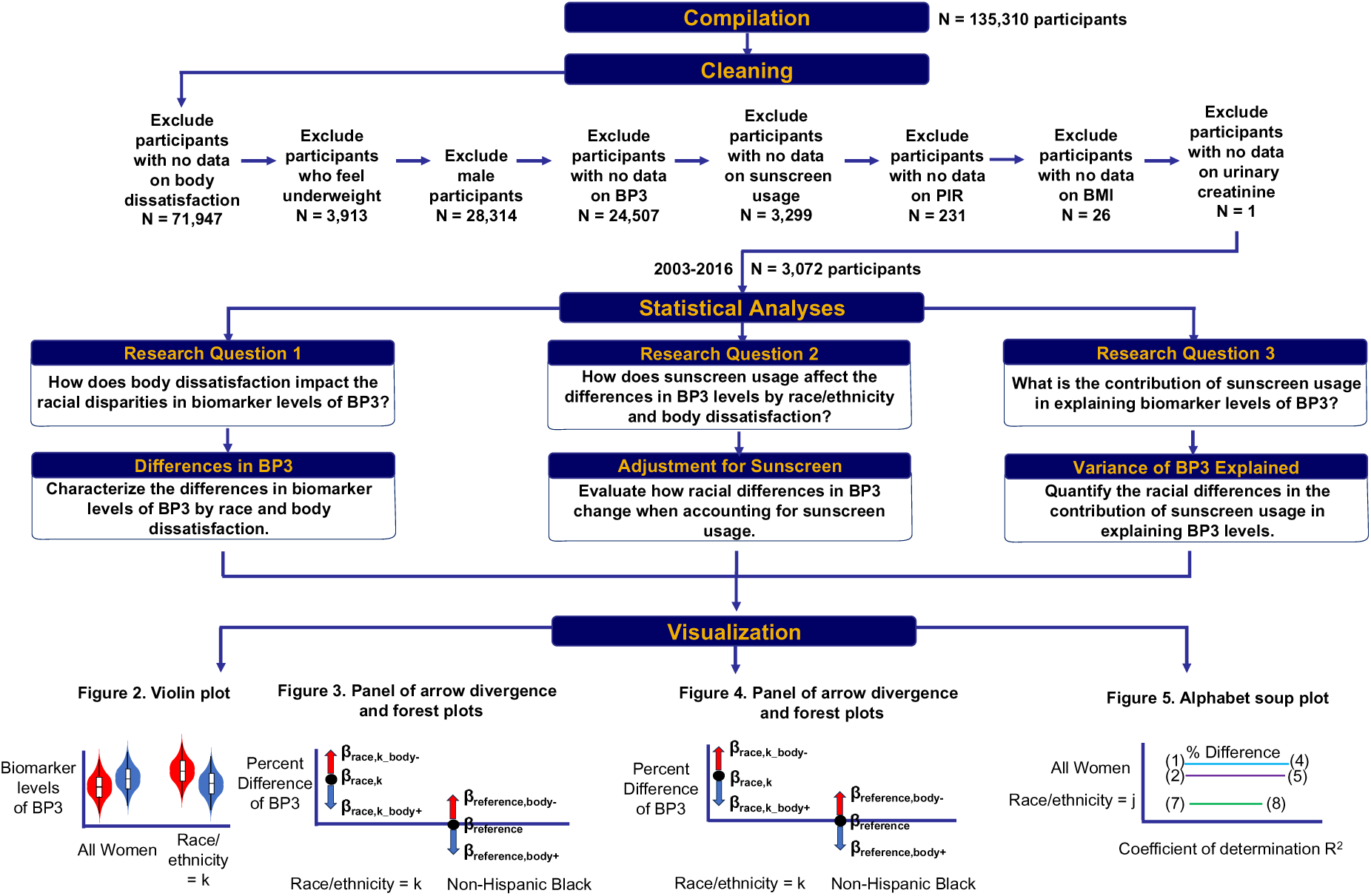
Schematic description of workflow on inclusion criteria of participants and of the statistical methods used to characterize the impact of body dissatisfaction on the racial disparities in biomarker levels of BP3.

### 2.2 Body dissatisfaction

We defined body dissatisfaction using overweight preoccupation. We overweight preoccupation as a surrogate variable for body dissatisfaction due to the high positive correlation between overweight preoccupation and other variables related to body dissatisfaction such as body image discrepancy, appearance importance, and appearance dissatisfaction^27,28^. We classified participants as having body dissatisfaction if they indicated feeling overweight in response to the question "How do you consider your weight?"

### 2.3 Statistical analysis

We conducted our analyses using R version 4.3.1. Our analytical code is publicly available online at https://github.com/vynguyen92/weight_perception_bp3. We accounted for the NHANES sampling design by applying the appropriate survey weights, strata, and primary sampling units in our regression models to ensure that estimates are generalizable to the US population. We used the R *survey* package. The general steps for statistical analyses are delineated in Figure 1 with more details delineated in Figure S1.

#### 2.3.1 Collated survey weights for urinary BP3

We used collated NHANES survey weights for urinary BP3 developed in our previous studies to calculate population statistics for the US population^25,26^. To analyze urinary BP3 across the study period of 2003-2016, we used specific survey weights variables for each NHANES cycle: WTSC2YR (2003-2004), WTSB2YR (2005-2010), WTSA2YR (2011-2012), and WTSB2YR (2015-2016). To simplify the analysis across multiple NHANES cycles, we consolidated all these weight variables for urinary BP3 ("URXBP3”) in one variable labeled “WT_URXBP3”. The survey weights in a given cycle represents the entire US population at that time, meaning the total of all survey weights equals the number of US residents. Since we combined data from 7 NHANES cycles, we adjusted the survey weights to represent a single US population rather than 7. We did this by dividing the survey weights by the number of NHANES cycles, which 7 for the case of urinary BP3. We calculated the populations statistics using the R *survey* and display the statistics using the *gtsummary* package.

#### 2.3.2 Impact of body dissatisfaction on racial disparities in BP3

We evaluated how body dissatisfaction changes the racial differences in biomarker levels of BP3 by running multivariate regression models. The outcome variable is the log10-transformed urinary concentrations of BP3, which we used as a surrogate for using personal care products. The covariates include age (continuous), NHANES cycles (continuous), urinary creatinine (continuous), BMI (continuous), and poverty income ratio (continuous). We included age to adjust for differences in exposures to BP3 across the lifespan. We included NHANES cycles to adjust for changes in BP3 levels over time. The poverty income ratio (PIR) is a ratio of the household income and poverty threshold based on the participant’s family size. We used PIR as a surrogate variable for socioeconomic status. We included creatinine levels to adjust for differences in urine dilution and urinary flow^29^ (Barr et al., 2005). As higher BMI is associated with greater body dissatisfaction^30–32^, we adjusted for BMI to examine the effect of weight perception and race on BP3 irrespective of the participant’s actual body size. We further examined whether there is a difference in the trend of BMI in participants with vs. without overweight preoccupation by conducting a linear regression model with BMI as the outcome variable, the main predictor as weight perception, and the covariates as age and poverty income ratio. We conducted this in the entire NHANES women population as well by race/ethnicity.

To examine the impact of body dissatisfaction on the racial disparities of BP3, we ran two sets of models: one where the main predictor is race (Equation 1), while the other where the main predictor is the pairwise combination of race and body dissatisfaction (Equation 2).

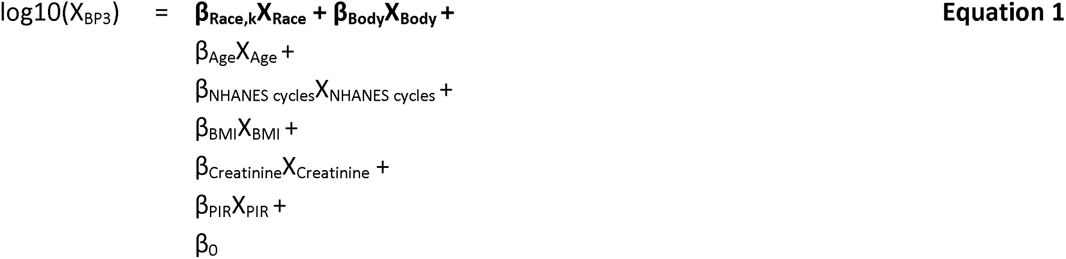

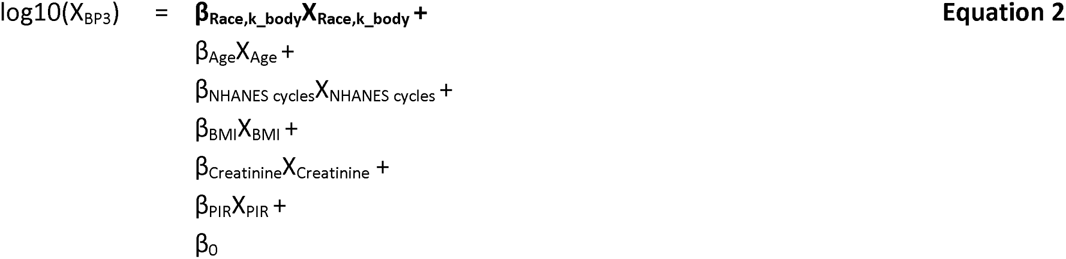

Here, *X*_BP3_ is the biomarker concentration for BP3, where k IZ {Mexican American, Other *Hispanic, non-Hispanic Asian, non-Hispanic White, Other Race - Including Multi-Racial}.* The reference group is the non-Hispanic Black women to ensure ease of comparison. We chose non-Hispanic Black women as the reference group due to how this group, on average, has the lowest biomarker levels of BP3. An example of a pairwise combination of race and body dissatisfaction is “Other Hispanic women who perceived themselves to be at the right weight” or “Other Hispanic women who perceived themselves to be overweight”. For a given race/ethnicity (*k*), there is a regression coefficient (*β*_race,k_) for comparing the race/ethnicity to the reference and two regression coefficients for comparing the groups "perceived at the right weight" (*β*_race,k+_) and "perceived as overweight" (*β*_race,k-_) to the reference. For ease of interpretation, we converted these coefficients to percent differences [10^β^ - 1] x 100. Moreover, we cannot use Equations 1 and 2 to extract the *β*_race,k+_ and *β*_race,k-_ for the reference group of Non-Hispanic Black women. Thus, we conducted a stratified analysis in non-Hispanic Black women where we compared both “non-Hispanic Black women who perceived themselves to be at the right weight” and “non-Hispanic Black women who perceived themselves to be overweight” to the same reference group of all non-Hispanic Black women (Equation 3).

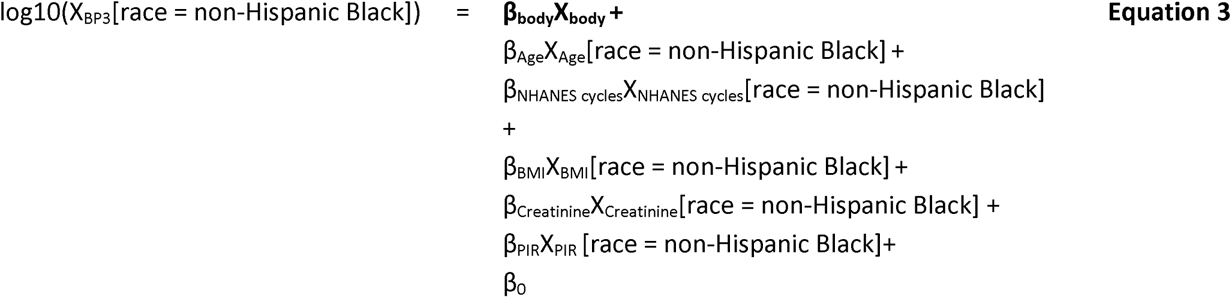

#### 2.3.3 Adjusting for sunscreen usage

We conducted a sensitivity analysis to examine how sunscreen usages changes the impact of body dissatisfaction on the racial differences in biomarker levels of BP3. Thus, to account for the participants’ usage of sunscreen, we additionally adjusted for sunscreen usage (ordinal) in the multivariate regression models for all NHANES women (Equation 4 and 5) and non-Hispanic Black women (Equation 6).

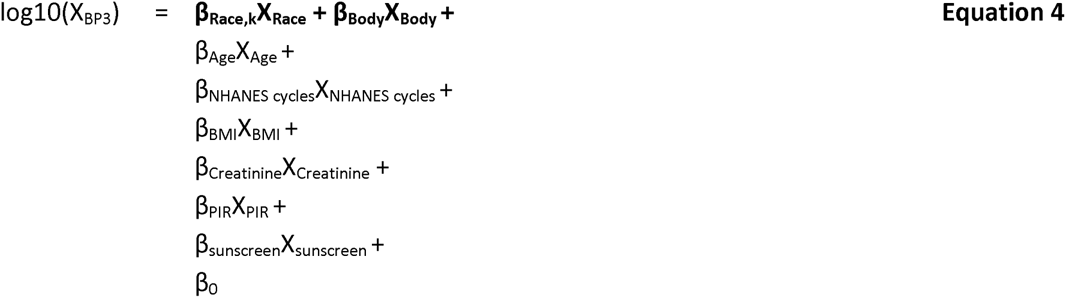

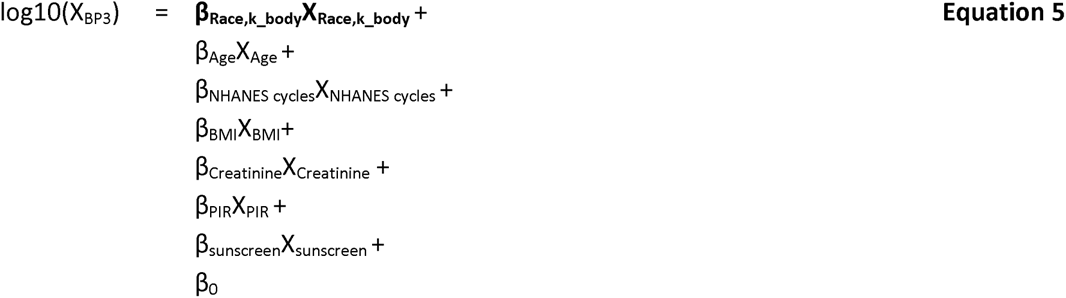

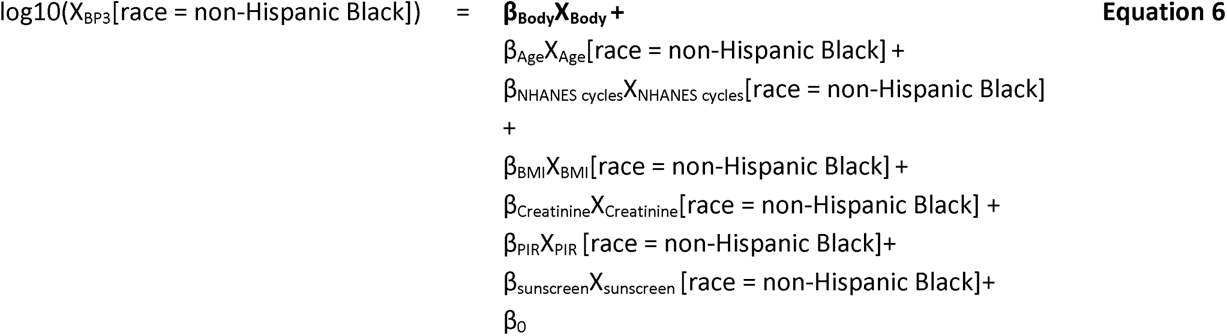

We used the questionnaire “Use sunscreen?” from the NHANES questionnaire on dermatology as the variable to represent sunscreen usage. The categories were encoded as the following: 1 (always), 2 (most of the time), 3 (sometimes), 4 (rarely), and 5 (never). To facilitate ease of interpretation for the higher values to mean more usage of sunscreen, we reversed the values. We examine the differences in sunscreen usage across the race/ethnicity by conducting an ordinal regression model with the outcome variable as sunscreen usage and the main predictors as race/ethnicity with Non-Hispanic Black as the reference group. The covariates are PIR and age. Additionally, we assessed how sunscreen usage differs by race and weight perception categories by running a similar ordinal regression model with the main predictor as the combination of race/ethnicity and weight perception. To examine how sunscreen usage differs by weight perception in the reference group of Non-Hispanic Black women, we stratified by race/ethnicity.

#### 2.3.4 Contribution of sunscreen usage in explaining BP3 levels

We are interested in understanding how sunscreen usage explains the variation in BP3 and how this varies among different races/ethnicities. Thus, we conducted a sensitivity analysis to examine how the coefficient of variation (R^2^) changes with and without adjusting for sunscreen usage in the NHANES women population (Equations 1-2, 4-5) and in each race/ethnicity (Equations 7-8), where j IZ {Mexican American, Other Hispanic, non-Hispanic *Asian, non-Hispanic White, non-Hispanic Black, Other Race - Including Multi-Racial}*.

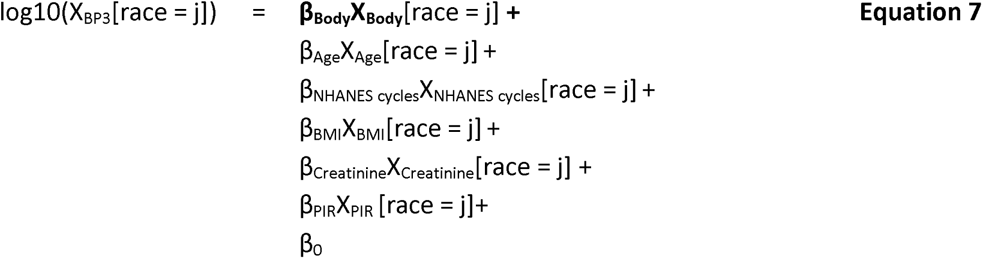

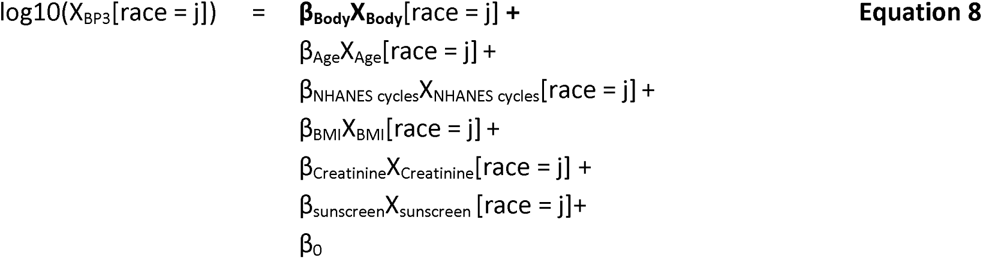

## Results

Table 1 displays the population characteristics of the study population. We leveraged 2003-2016 national biomonitoring data on 3,072 NHANES female participants. These participants have data on urinary creatinine, BMI, poverty income ratio, urinary BP3, body dissatisfaction, and sunscreen usage. The participants self-reported their race/ethnicity, which the responses include Non-Hispanic White (1,269 subjects), Non-Hispanic Black (703 subjects), Mexican American (479 subjects), Other-Hispanic (265 subjects), Non-Hispanic Asian (210 subjects), and Other Race, including multi-racial (146 subjects). The average age of the participants is 39.5 years old with the Non-Hispanic White group as the oldest at 40.3 years and the Mexican American group as the youngest at 36.9 years. The range of age is 20-59 years. The average urinary creatinine levels of all participants is 110.5 mg/dL with the highest concentrations found in Mexican Americans at 111.2 mg/dL and lowest concentrations found in Non-Hispanic Asians at 93.6 mg/dL. The average BMI for all participants is 29.0 kg/m^2^ with the highest BMI found in Mexican Americans at 30.5 kg/m^2^ and lowest found in Non-Hispanic Asians at 24.2 kg/m^2^. The average BMI for Non-Hispanic Asian was greater than 1 standard deviation from the mean of all NHANES participants. All racial/ethnic groups have poverty income ratio (PIR) within one standard deviation of the mean PIR for all NHANES participants.

**Table 1.** Population statistics of NHANES women. For the continuous variables, the mean and standard deviations are calculated. For the categorical variables, the participant counts are not survey weighted, while the percentages were survey weighted.

| Characteristics | All NHANES Women<br>N = 3072<sup>†</sup> | Non-Hispanic White<br>N = 1269<sup>†</sup> | Mexican American<br>N = 479<sup>†</sup> | Other Hispanic<br>N = 265<sup>†</sup> | Non-Hispanic Black<br>N = 703<sup>†</sup> | Non-Hispanic Asian<br>N = 210<sup>†</sup> | Other Race - Including Multi-Racial<br>N = 146<sup>†</sup> |
| --- | --- | --- | --- | --- | --- | --- | --- |
| Age (years) | 39.5 (11.3) | 40.3 (11.3) | 36.9 (10.5) | 37.1 (10.9) | 38.6 (11.4) | 37.6 (10.5) | 39.0 (11.5) |
| Urinary creatinine (mg/dL) | 110.5 (77.1) | 101.4 (73.1) | 111.2 (66.8) | 109.8 (71.4) | 160.3 (87.0) | 93.6 (64.1) | 106.4 (79.9) |
| Body Mass Index (kg/m**2) | 29.0 (7.6) | 28.3 (7.4) | 30.5 (6.8) | 30.1 (6.5) | 32.6 (8.9) | 24.2 (4.4) | 27.4 (7.3) |
| Poverty income ratio (-) | 2.97 (1.68) | 3.32 (1.61) | 1.91 (1.42) | 2.32 (1.55) | 2.16 (1.60) | 3.22 (1.72) | 2.93 (1.55) |
| Urinary Benzophenone-3 (ng/mL) | 421.1 (1,581.9) | 423.6 (1,468.5) | 306.4 (1,040.2) | 496.5 (1,628.1) | 305.3 (1,454.3) | 380.9 (1,310.9) | 829.3 (3,290.0) |
| Weight perception |  |  |  |  |  |  |  |
| perceived about the right weight | 1,114 (35%) | 456 (34%) | 158 (32%) | 77 (27%) | 238 (33%) | 127 (60%) | 58 (43%) |
| perceived overweight | 1,958 (65%) | 813 (66%) | 321 (68%) | 188 (73%) | 465 (67%) | 83 (40%) | 88 (57%) |
| Sunscreen usage |  |  |  |  |  |  |  |
| always | 519 (19%) | 256 (22%) | 68 (15%) | 69 (24%) | 51 (7.3%) | 51 (25%) | 24 (12%) |
| most of the time | 475 (20%) | 302 (25%) | 51 (11%) | 29 (14%) | 28 (3.4%) | 39 (18%) | 26 (19%) |
| sometimes | 622 (23%) | 309 (25%) | 103 (23%) | 61 (23%) | 75 (11%) | 43 (22%) | 31 (27%) |
| rarely | 402 (13%) | 170 (13%) | 65 (13%) | 32 (11%) | 93 (13%) | 24 (10%) | 18 (13%) |
| never | 1,054 (25%) | 232 (15%) | 192 (38%) | 74 (29%) | 456 (65%) | 53 (24%) | 47 (29%) |

<sup>†</sup> Mean (SD); n (unweighted) (%)

Mexican Americans have the lowest PIR (1.91), indicating the lowest socioeconomic status among all the racial/ethnic groups, while Non-Hispanic Asians have the highest PIR (3.22), indicating the highest socioeconomic status. The average urinary level of BP3 for all participants is 421.1 ng/mL with the highest concentrations found in the Other Race group at 829.3 ng/mL and the lowest concentrations found in the Non-Hispanic Black at 305.3 ng/mL. Most participants perceived themselves as overweight (65%), which is also the trend found in the racial/ethnic groups except for the Non-Hispanic Asian group. In contrast, a smaller portion of Non-Hispanic Asians perceived themselves as overweight (40%). The majority of all NHANES participants (75%) and participants of various races/ethnicities (62%-85%) have used sunscreen, except for Non-Hispanic Black participants, of whom only 35% have used sunscreen (Table 1).

### 3.1 Impact of body dissatisfaction on racial disparities in BP3

Figure 2 is a violin plot comparing the creatinine-normalized levels of BP3 between women who perceived themselves as overweight to those who perceived themselves at the right weight. The comparisons are among all NHANES women in the study and stratified by race/ethnicity. Non-Hispanic White women, who perceive themselves as the right weight, have significantly higher BP3 levels (p < 0.05) than those who perceive themselves as overweight. By contrast, Non-Hispanic Black and Other Hispanic women, who perceive themselves as the right weight, have lower BP3 levels (p > 0.05) than those who perceive themselves as overweight.

**Figure 2.**
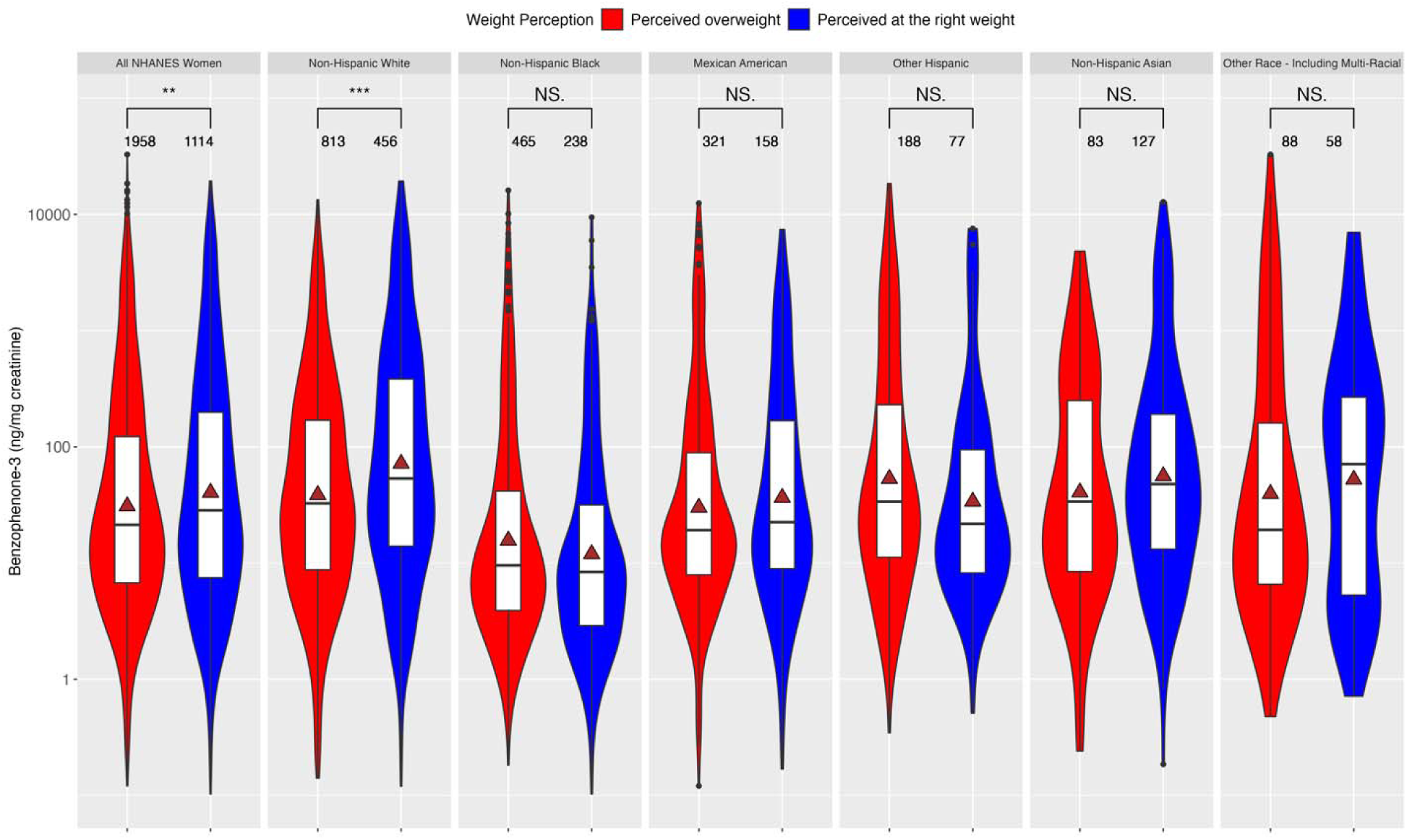
Violin plot of creatinine-normalized levels of BP3 stratified by race and weight perception for all NHANES women. The gray triangle represents the mean biomarker concentration of BP3. The number of asterisks indicates the statistical significance of the differences in urinary levels of BP3: *(p-value ⍰ [0.01,0.05]), **(p-value ⍰ [0.001,0.01]), and ***(p-value ≤ 0.001).

Table S1 shows how body weight perception is associated with BMI for all NHANES women and by each race/ethnicity. We conducted this analysis to examine differences in the BMI by weight perception. On average, those who perceived themselves as overweight had BMIs that were 5.72-9.64 kg/m² higher than those who perceived themselves at the right weight. These differences were significant with p-values ranging from 7.24E-222 to 9.81E-13. Thus, we adjusted all models for BMI to account for it as a confounder and to examine the difference in BP3 levels between those with and without body dissatisfaction, beyond their actual weight.

Figure 3A is an arrow divergence plot that shows the racial differences in urinary levels of BP3 (black dot) and how these racial differences change when a group perceives themselves as overweight (red arrow) vs. at the right weight (blue arrow) for all NHANES participants. We tested the hypothesis that body dissatisfaction would be associated with higher BP3 levels in minority groups. The fold differences are adjusted for age, NHANES cycle, BMI, and poverty income ratio and are tabulated in Table S2. The black dot represents the fold differences in BP3 between a given race/ethnicity and the reference group of Non-Hispanic Black women.

**Figure 3.**
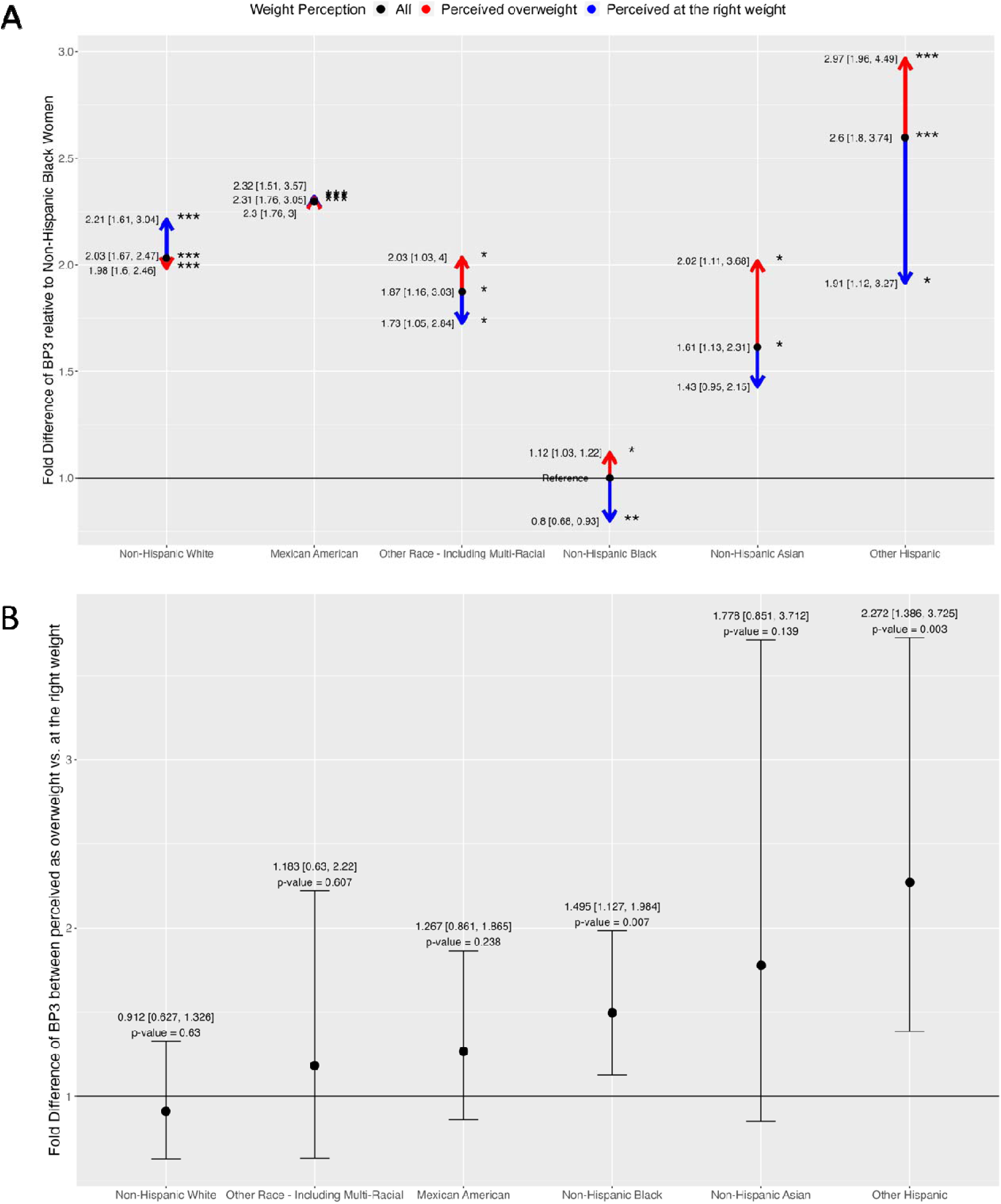
Panel plot of fold differences of BP3, adjusted for age, NHANES cycle, body mass index (BMI), and poverty income ratio (PIR). The number of asterisks indicates the statistical significance of the fold differences: *(p-value ⍰ [0.01,0.05]), **(p-value ⍰ [0.001,0.01]), and ***(p-value ≤ 0.001). A) Arrow divergence plot of adjusted fold differences of BP3 in all NHANES women. The reference groups are non-Hispanic Black women for the adult participants. B) Forest plot adjusted fold differences of BP3 between those who perceived themselves as overweight vs. at the right weight, stratified by race/ethnicity.

Compared to Non-Hispanic Black women, BP3 levels were 2.59 times higher in Other Hispanic women (p-value = 1.83E-06), 2.30 times higher in Mexican American women (p-value = 3.79E-08), 2.03 times higher in Non-Hispanic White women (p-value = 4.25E-10), 1.87 times higher in Other Race women (p-value = 0.012), and 1.61 times higher in Non-Hispanic Asian women (p-value = 0.011). The red arrow represents the fold difference in BP3 between a racial/ethnic group with body dissatisfaction (e.g., perceived as overweight) and the reference group of Non-Hispanic Black women. For example, BP3 levels of Other Hispanics who perceive themselves as overweight were 2.97 times higher than the levels in Non-Hispanic Blacks. Notably, this difference is greater than the difference observed when comparing the Other Hispanic group to Non-Hispanic Blacks at a fold difference of 2.59. On the other hand, the blue arrow represents the fold difference between a racial/ethnic group who perceived themselves at the right weight and the same reference group. For example, BP3 levels of Other Hispanics who perceive themselves at the right weight were 1.91 times higher than the levels in Non-Hispanic Blacks.

Notably, this difference is lower than the difference observed when comparing the Other Hispanic group to Non-Hispanic Blacks. More specifically, the fold difference in BP3 levels between Non-Hispanic Asian vs. Black women increased from 1.61 times to 2.02 times when considering Non-Hispanic Asian women who perceived themselves as overweight. Between Other Race and Non-Hispanic Black women, BP3 levels significantly increased from 1.87 to 2.03 times higher when considering Other Race women with overweight perception. BP3 levels in Non-Hispanic Black women with overweight perception are 1.12 times higher compared to the entire group of Non-Hispanic Black women, while the BP3 levels are 0.8 times lower in those perceiving themselves at the right weight. These trends are illustrated by the red arrows pointing upward, the blue arrows pointing downward, and both arrows diverging from the black dot. By contrast, only in Non-Hispanic White women did BP3 levels decrease from 2.03 times to 1.98 times when comparing Non-Hispanic White women with overweight perception to Non-Hispanic Black women. Overall, in the Other Hispanic, Non-Hispanic Asian, Other Race, and Non-Hispanic Black groups, body dissatisfaction widens the racial differences in BP3 levels: higher levels are shown in those with overweight perception, and lower levels are shown in those who perceived themselves at the right weight. The exception is found in Non-Hispanic White women.

Figure 3B is a forest plot showing the fold difference in BP3 levels between women who perceived themselves as overweight vs. those who perceived themselves at the right weight for each race/ethnicity. The fold differences are adjusted for age, NHANES cycle, BMI, and poverty income ratio and are tabulated in Table S3. BP3 levels were higher in women with overweight perception compared to their counterparts who perceived themselves at the right weight: 2.27 times in Other Hispanic, 1.77 times in Non-Hispanic Asian, 1.49 times in Non-Hispanic Black, 1.27 times in Mexican American, 1.18 times in Other Race, and 0.91 times in Non-Hispanic White women. The differences are significant in Other Hispanic (p-value = 0.003) and Non-Hispanic Black (p-value = 0.007).

### 3.2 Adjusting for sunscreen usage

Figure 4A is an arrow divergence plot that shows the racial differences in BP3 and how these racial differences change when considering perception as overweight and perception at the right weight for all NHANEs participants, additionally adjusting for sunscreen usage. The fold differences are tabulated in Table S4. As BP3 is a common ingredient in sunscreen, we examined how the associations between BP3 and weight perception changed when adjusting for sunscreen usage, in addition to adjusting for age, NHANES cycle, BMI, and PIR. The racial differences in BP3 levels attenuated when adjusting for sunscreen usage. Without this adjustment, the fold differences ranged from 1.61-2.59, but with the adjustment, the range decreased to 1.09-1.59. Relative to Non-Hispanic Black women, BP3 levels remained significantly higher in Mexican Americans (1.59, p-value = 0.00043) and Other Hispanic women (1.56, p-value = 0.02). The racial differences in BP3 levels by weight perception also attenuated. The ranges changed from 0.8-2.97 without adjusting for sunscreen to 0.85-1.69 when adjusting for sunscreen usage. Relative to Non-Hispanic Black women, urinary levels of BP3 are still significantly higher in Mexican American women (1.64, p-value = 0.00072) and Other Hispanic women (1.69, p-value = 0.014) with overweight perception. For all races/ethnicities except for Non-Hispanic White women, body dissatisfaction still widens the racial differences in BP3 levels. Higher levels are shown in those with overweight perception (i.e., red arrows pointing upward) and lower levels are shown in those who perceived themselves at the right weight (i.e., blue arrows pointing downward). Overall, while adjusting for sunscreen usage partially explains the racial differences in BP3 levels by weight perception, the significant differences still remain.

**Figure 4.**
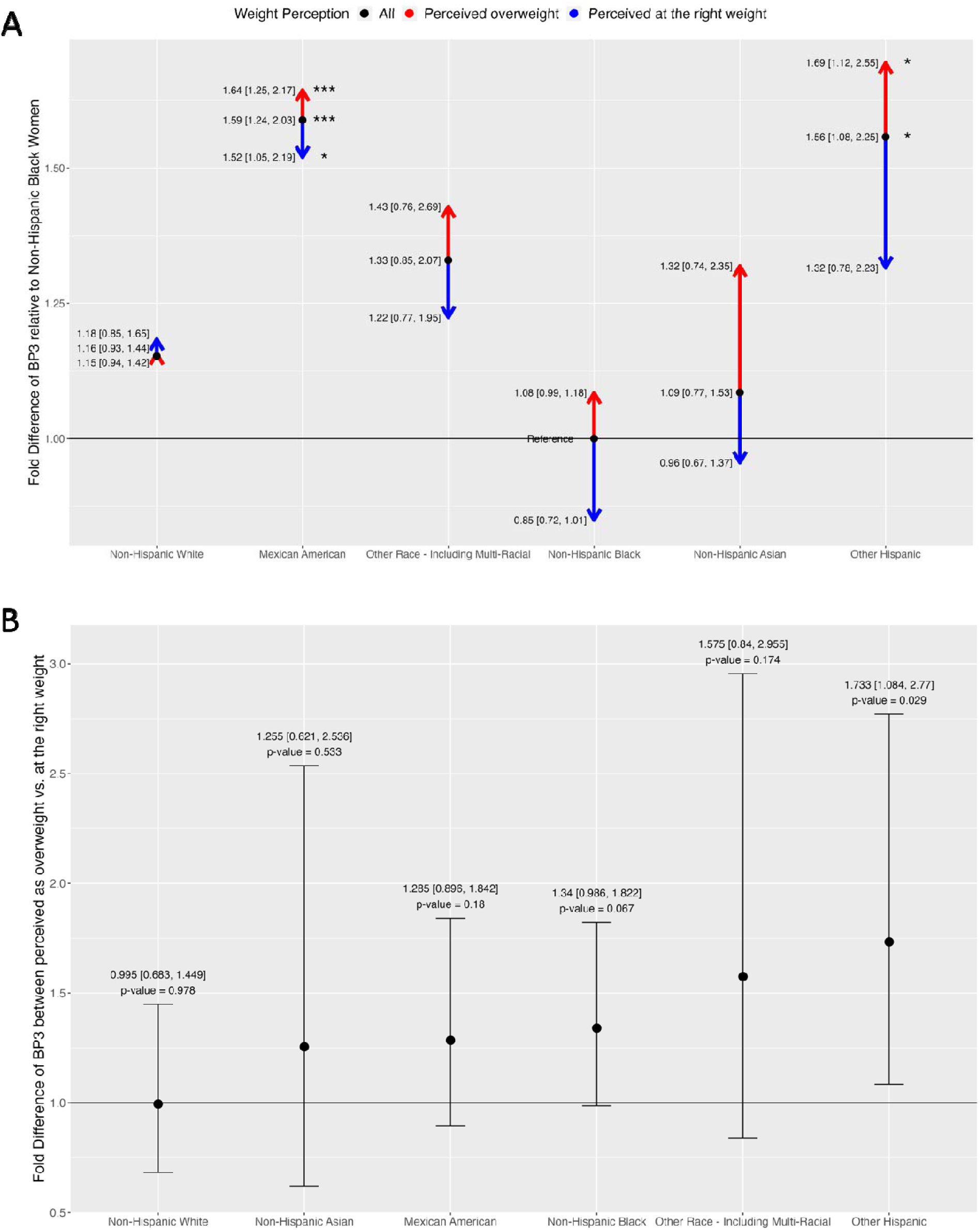
Panel plot of fold differences of BP3, additionally adjusted for sunscreen. All results are adjusted for age, NHANES cycle, body mass index (BMI), poverty income ratio (PIR) and sunscreen usage. The number of asterisks indicates the statistical significance of the fold differences: *(p-value ⍰ [0.01,0.05]), **(p-value ⍰ [0.001,0.01]), and ***(p-value ≤ 0.001). A) Arrow divergence plot of adjusted fold differences of BP3 in all NHANES women. The reference groups are non-Hispanic Black women for the adult participants. B) Forest plot adjusted fold differences of BP3 between those who perceived themselves as overweight vs. at the right weight, stratified by race/ethnicity.

Figure 4B is a forest plot showing the fold difference of BP3 levels between women who perceived themselves as overweight vs. those who perceived themselves at the right weight for each race/ethnicity, additionally adjusting for sunscreen usage. The fold differences are tabulated in Table S5. The differences attenuated when adjusting for sunscreen usage. The ranges changed from 0.91-2.27 without adjusting for sunscreen to 0.99-1.73 when adjusting for sunscreen usage. As before, urinary levels of BP3 were higher in minority women who perceived themselves as overweight, ranging from 1.25-1.73, but was only significant in Other-Hispanic Women (1.73, p-value = 0.029).

Table S6 shows the odd ratios of frequently using sunscreen (always using sunscreen vs. most of the time, sometimes, rarely, or never) between a given race/ethnicity and the reference group of Non-Hispanic Black women, while Table S7 shows how these odd ratios change when considering weight perception. We evaluated the associations between sunscreen usage and race/ethnicity. Non-Hispanic White, Other Hispanic, Non-Hispanic Asian, Other Race, and Mexican American women are 5.49, 5.11, 4.99, 3.81, and 3.50 times as likely as Non-Hispanic Black women to frequently wear sunscreen. Then we evaluated how these associations change when considering weight perception. For example, Non-Hispanic White women who perceived themselves at the right weight were 6.31 times as likely to frequently use sunscreen as Non-Hispanic Black women (p-value = 1.28E-228). The frequency is lower in their counterparts with overweight perception. Non-Hispanic White women who perceived themselves as overweight were 5.13 times more likely to frequently use sunscreen as Non-Hispanic Black women (p-value = 2.35E-238). This pattern is similar for Non-Hispanic Asian (6.21 vs. 4.06), Other Hispanic (5.17 vs. 5.10), Other Race (4.07 vs. 3.71), and Mexican American (3.57 vs. 3.46). Overall, for all races/ethnicities, participants who perceived themselves as overweight are less likely to wear sunscreen than those who perceived themselves at the right weight.

Table S8 shows the odd ratios of frequently using sunscreen between women with and without overweight perception in each race/ethnicity. In addition to running models on all races/ethnicities together, we also examined the association between sunscreen usage and weight perception in each race/ethnicity individually. This allows us to view the association between sunscreen and weight perception in Non-Hispanic Black women, which was used as the reference group in the previous models. Non-Hispanic Black women are 6% (p-value = 0.5) more likely to wear sunscreen when they perceive themselves as overweight. In contrast, Non-Hispanic Asian, Non-Hispanic White, all NHANES participants, Other Race, Other Hispanic, and Mexican American women are 32.2%, 21%, 15.2%, 7.9%, 6.6%, and 2% less likely to wear sunscreen when they perceive themselves as overweight, respectively. The differences are significant in Non-Hispanic Asian (p-value = 8.93E-04), Non-Hispanic White (p-value = 6.95E-06), and all NHANES participants (p-value = 8.42E-07).

### 3.3 Contribution of sunscreen usage in explaining BP3 levels

Figure 5 shows the coefficient of variation (R^2^) between models without and with adjusting for sunscreen usage for all NHANES participants and in each race/ethnicity individually. The R^2^ quantified the contribution of sunscreen usage to urinary BP3 levels and are tabulated in Table S9. Sunscreen usage increased the R² by 11.7% in Non-Hispanic Asians, 8.8% in Mexican Americans, 8% in Other Hispanics, 7.4% in Non-Hispanic Blacks, 6.5% in Non-Hispanic Whites, and 20.7% in women of Other Race. For all NHANES participants, sunscreen usage increased the R^2^ by 7.7% and 7.8% in two different models: one model with race/ethnicity and body dissatisfaction as two separate variables and another with the combination of these two variables. These results show that sunscreen usage is a small but significant source of BP3 exposures in women. Additionally, there are other personal care products that are potential sources of BP3 exposures in women.

**Figure 5.**
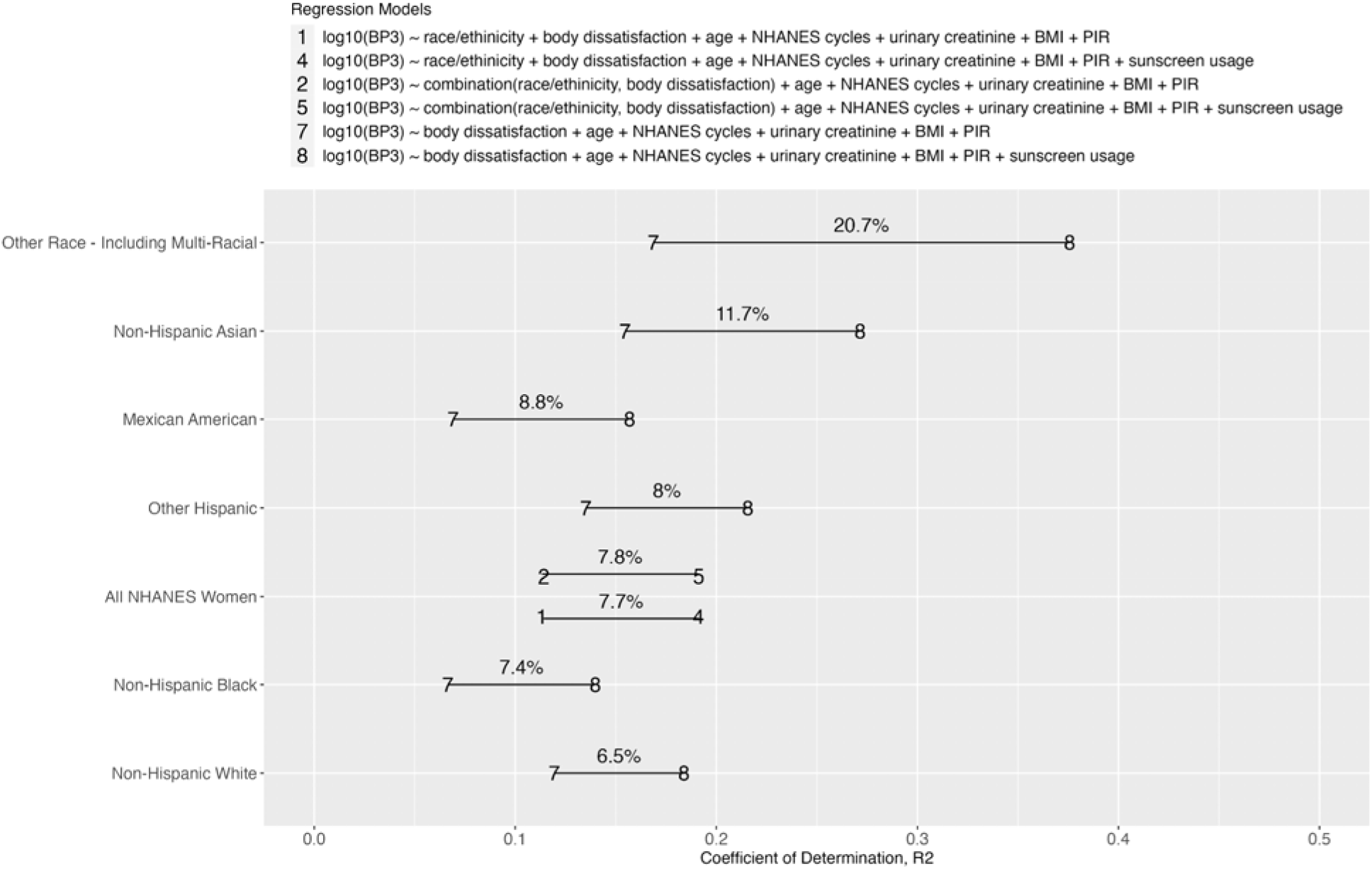
Alphabet soup plot of coefficient of determination, R^2^ by different set of adjustment for confounders in all NHANES women and by race/ethnicity. The percentages are the difference in R between without vs. with adjusting for sunscreen usage. These percentages represent the contribution of sunscreen usage in explaining biomarker levels of BP3. Models (1)-(2) and (4)-(5) are conducted on the NHANES women population, while Models (7) and (8) are conducted for each race.

## 4. Discussion

In this study, our findings highlight the potential relationship between body dissatisfaction and BP3 across race/ethnicity. These differences are still prominent even when accounting for differences in sunscreen usage. From the analysis on the differences of BP3 by race/ethnicity and body dissatisfaction, we found that within the same minority group, adult women perceiving themselves as overweight have higher BP3 levels than those perceiving themselves at the right weight. The opposite is true for non-Hispanic White adult women.

Sunscreen usage attenuates these differences, but even after adjusting for sunscreen usage, the disparities in BP3 between perceiving overweight vs. at the right weight persists for minority adults, but not for non-Hispanic White adults. Quantifying the contribution of sunscreen usage in explaining BP3 levels, we observed that the highest to lowest proportion of BP3 explained by sunscreen usage is in the following order: Other Race, Non-Hispanic Asian, Mexican American, Other Hispanic, All NHANES Adult Women, Non-Hispanic Black, and Non-Hispanic White. Overall, our findings show that minority women with body dissatisfaction show elevated BP3 exposure independent of their sunscreen usage, implying that their elevated exposures may stem from using other personal care and consumer products.

Prior studies observed that the non-Hispanic White women have the highest biomarker levels of BP3 and attribute such findings to elevated usage of sunscreen in non-Hispanic White women compared to others^14,25,33^. We confirmed non-Hispanic White women on average used the most sunscreen and had the highest levels of BP3. However, after we accounted for sunscreen use, non-Hispanic White women had lower BP3 levels than Mexican Americans, Other Hispanic, and Other Race including Multi-Racial women. Our findings in the adult population imply that while sunscreen usage contributes to higher BP3 levels, it is not the sole factor explaining racial differences in BP3 levels. Moreover, after accounting for sunscreen usage, we observed that the racial differences in BP3 decreased by almost half but still persist. Our discovery has implications for other chemicals highly correlated with elevated usage of sunscreen and other personal care products such as phthalates, triclosan, and parabens. It sheds light on how accounting for sunscreen usage may attenuates the racial disparities in these chemicals, but the disparities may still persist.

Furthermore, we showed that sunscreen use only explains a limited proportion of the variability of BP3 biomarker levels with the R2 ranges from 6.5% to 20.7%. Our findings corroborate the findings of Zamoiski et al., 2015, where R2 ≤ 0.14^34^. Notably, sunscreen explains the smallest variability in BP3 levels among non-Hispanic Whites (6.5%), compared to 7.4% to 20.7% among other minority women. This suggests that factors beyond sunscreen contribute significantly to BP3 levels, likely including behaviors related to personal care and consumer products.

Observational studies have found that increased exposures to BP3 is associated with using personal care and consumer products such as shaving cream^35^, mouthwash, dental rinse^36^, eye make-up, hair oil^37^, hair leave-in styling, shampoo^38^, hand or body lotion, face cream, lip balm^39,40^, and plastic water bottle^40^. Usage patterns of the aforementioned products vary by race. For instance, among Californian women, Latina women used cosmetics like eye make-up and all lip products more frequently relative to other races/ethnicities. Black women used hair oil and body lotion. Vietnamese women use facial cleansing products. White women use eyeshadow, lip balm/chapstick, and face cream^41,42^. A limitation of our study was that NHANES did not have data on usage frequency of other personal care and consumer products. Thus, future work can apply such questionnaires to gather data on usage patterns at the national levels. These data would enable future research to explore how other personal care and consumer products contribute to explaining the differences in BP3 levels by race/ethnicity and body dissatisfaction.

We used overweight preoccupation as a surrogate for body dissatisfaction, given its high positive correlation with other variables related to body dissatisfaction^27,28^. Our observation revealed that individuals with overweight preoccupation generally exhibit higher BMI compared to those perceiving themselves at the right weight. Thus, we included weight perception and BMI into the same regression model to represent the participant’s dissatisfaction with their body beyond their actual weight. Furthermore, our analysis uncovered intriguing patterns with weight perception versus BMI regarding biomarker levels of BP3. Overweight preoccupation was associated with higher biomarker levels of BP3. In contrast, higher BMI (i.e., being overweight) was associated with lower BP3 levels, suggesting a decrease in urinary levels of BP3 with increasing BMI. The contrasting patterns in how weight perception and actual weight relate to BP3 levels further augments how weight perception goes beyond just weight itself in capturing participants’ body dissatisfaction. Additionally, our examination of sunscreen usage and weight perception lent further support to overweight preoccupation as a proxy for body dissatisfaction. Participants, who perceived themselves at the right weight, use sunscreen more often than those who perceived themselves to be overweight. This pattern holds true across the NHANES women population and by race/ethnicity, except for non-Hispanic Black women.

These results align with existing research showing that people satisfied with their bodies tend to use sunscreen more often^43,44^. However, these findings contrast with our observation that minority women who perceived themselves to be at the right weight tend to have higher levels of BP3. This emphasizes that the higher BP3 exposure in minority women is not mainly due to sunscreen usage but to using other personal care products. Ideally, we would prefer to use the gold standard metrics of body dissatisfaction such as Eating Disorder Inventory-Body Dissatisfaction subscale (EDI-BD)^45^ and Multidimensional Body-Self Relations Questionnaire-Appearance Evaluation subscale (MBSRQ-AE)^46^. The EDI-BD and the MBSRQ-AE subscales are scales designed to measure dissatisfaction for specific body sites or for a person’s overall appearance, respectively. Incorporating these scales into future studies and national surveys, such as NHANES, could enhance our understanding of the impact of body dissatisfaction driving elevated chemical exposures, particularly in minority populations.

Targeted marketing aimed at women of color could encourages them to purchase personal care products to adhere to predominant White beauty standards^6^. These standards typically emphasize characteristics such as straight hair and lighter skin tone, reflecting deep-rooted hair discrimination, texturism, and colorism within society. Texturism is the preference towards straight hair and the discrimination against individuals with kinkiers, tighter-curled hair textures^47^. Colorism is characterized by preferential treatment towards individuals with lighter skin tones and bias against those with darker skin tones^48,49^. Despite such discrimination and pressure on women of color, minority women report slightly better body image than non-Hispanic White women. For instance, in a body image study in the US, they tend to have lower preoccupation with being overweight and higher satisfaction with their appearance^28^. Despite having slightly better body image, minority women still engage in using personal care products due to societal pressures and concerns about appearance. In a North and South Manhattan survey, a majority of minority women (61% and 57%) report using chemical straightening and skin lightening products to conform to beauty standards associated with straight hair and lighter skin, respectively^7^. Additionally, survey data on American college women in southeast Texas reveals that appearance dissatisfaction prompts 37% of them to begin using makeup^50^.

Consequently, the pervasive pressure and concerns surrounding appearance likely drive the purchase and use of personal care products, inadvertently exposing individuals to potential toxicants. This likely explains why we observed higher BP3 exposures in minority women with body dissatisfaction vs. those without. But such reasons do not explain the minimal differences in BP3 between non-Hispanic White women with and without body dissatisfaction. Non-Hispanic White women may perceive themselves as already aligning closely with prevailing standards and thus may not feel the same pressure to purchase and use personal care products to achieve these standards. Additionally, the promotion of White-centric features by the beauty industry may also be another explanation^51^. Alternatively, non-Hispanic White women may adopt different behaviors, unrelated to using personal care products. Despite the similar prevalence of body dissatisfaction across different racial/ethnic groups, the differential impact of body dissatisfaction on toxicant exposures may highlights how beauty standards influence behaviors, leading to different exposure patterns and associated health risks.

Adhering to beauty standards may increase the purchase and usage of personal care products in different racial/ethnic groups. In a qualitative study on extracting themes from questionnaires related to beauty and body image, a group of African American college women frequently describe meeting these beauty standards in terms of sacrifice^52^. Specifically, some endure financial debt by prioritizing the beauty expense over food^52^. In a qualitative study, a group of African American women experience anxiety about their hair due to being perceived as unprofessional^53^. In a study of students of African descent, increased dissatisfaction with ethnicity-specific attributes such as natural hair texture is significantly associated with adding chemicals and color to one’s hair^54^. Several studies in Black and Asian Americans found that the internalization of White beauty ideals predicted skin tone dissatisfaction as well as elevated usage of skin lightening and bleaching^7,55–57^. Even more concerning, only a small fraction of users of skin lightening products are aware of the potential adverse effects^57^. Such findings emphasize the importance of educational programs about the ingredients in the personal care products and the potential adverse health effects of using such products. Given that these products can contain BP3, frequent use among African and Asian American women striving to adhere to beauty standards may explain the elevated BP3 levels observed in non-Hispanic Black and Asian women with body dissatisfaction. Such information will be key in developing intervention strategies aimed at helping vulnerable individuals build resiliency against internalizing harmful beauty standards.

We observed that the differences in BP3 levels between women with and without body dissatisfaction varies within the Latina population. The highest differences in BP3 levels were observed in Other Hispanic women, with a 73% increase in BP3 levels for women experiencing body dissatisfaction compared to their counterparts without body dissatisfaction. This suggests a propensity for Other Hispanic women with body dissatisfaction to use more personal care and consumer products containing BP3, compared to those without body dissatisfaction. In comparison, Mexican American women with body dissatisfaction have 25% higher levels of BP3 compared to those without. Such a difference in Mexican American is a third of the magnitude observed for Other Hispanic women. These variations may be associated with other factors such as the level of assimilation into US culture. For example, a study of 202 Latina women found that those who assimilated into US culture show no association with makeup usages. In contrast, those more aligned with their Latina culture tended to use more beauty products^58^.

Additionally, a study of 211 Mexican of Mexican American college women found that those, who identify more with their Mexican culture, do not internalize US beauty standards nor experience related body dissatisfaction^59^. Thus, a future direction can be to incorporate variables related to assimilation such as immigrant status to help explain such variations in chemical exposures among Latina subgroups.

Our analysis has several limitations. First, as NHANES is a cross-sectional survey, we cannot make claims on causal factors of chemical exposures. Second, while there is data on sunscreen usage, there is no questionnaire to ascertain usage of personal care and consumer products. We would ideally like to have seen how such racial disparities can be explained by usage of other personal care and consumer products to better understand the sources of BP3 exposures. Furthermore, such information would have been useful in understanding how body dissatisfaction impacts such product usages in a diverse population. Third, there was a low sample size of participants who have data on sunscreen usage, which led to low sample size for participants who have data for all key variables in the analysis. Future studies can consider alternative approaches, such as using machine learning to predict sunscreen usage from correlated chemical biomarkers impute participant sunscreen usage to increase the sample size of the study population. Finally, we only consider a single chemical, which is not representative of actual exposures to multiple chemicals and mixtures. Thus, future studies can expand upon our work and deploy statistical tools to study the impact of body dissatisfaction on multiple chemical exposures and mixtures.

## 5. Conclusions

Evaluating the impact of body dissatisfaction on toxicant exposures provides another key factor to identifying susceptible populations. Our study is the first application to integrate data on body dissatisfaction with chemical biomonitoring data in a nationally representative sample of the US population. This integration bridges the field of body image with environmental health to light the way for studying the role of body dissatisfaction as a driver of exposures to other chemicals. Furthermore, this integration enabled us to develop a visualization tool to show how the racial disparities in biomarker levels of BP3 further widens when considering body dissatisfaction. Using this tool, we determined that the differences in biomarker levels of BP3 are higher in minority women with body dissatisfaction compared to those without, but such differences in the non-Hispanic White women are minimal. These findings have implications to study how body dissatisfaction from adhering to beauty standards in different races/ethnicities may lead to increased exposure to BP3 and other environmental toxicants. This information can help prioritize personal care products used for adhering to beauty standards and explore the differences in product use and reasons for using such products across races/ethnicities. Such insights can help guide future epidemiologic studies and intervention to help build resilience against body dissatisfaction and prevent the effects of toxicant exposures from using risky personal care products.

## Author contributions

**Vy Kim Nguyen** : conceptualization, data curation, methodology, formal analysis, software, validation, visualization, writing - original draft, writing - review & editing

**Samuel Zimmerman**: conceptualization, methodology, formal analysis, software, validation, visualization, writing - original draft, writing - review & editing

**Justin Colacino**: methodology, writing - review & editing

**Olivier Jolliet**: methodology, writing - review & editing, resources

**Chirag J Patel**: supervision, methodology, writing - review & editing, funding acquisition

## Data availability

The datasets, metadata, and starter codes supporting this article are publicly available in figshare^60^ (https://doi.org/10.6084/m9.figshare.21743372.v7), Kaggle^61^ (https://doi.org/10.34740/KAGGLE/DSV/9019774), and Hugging Face^62^ (https://huggingface.co/datasets/nguyenvy/cleaned_nhanes_1988_2018). Our analytical code is publicly available online at https://github.com/vynguyen92/weight_perception_bp3.

