## Supplemental Figures for "Body dissatisfaction widens the racial disparities of Benzophenone-3, a chemical biomarker of personal care and consumer product usage"

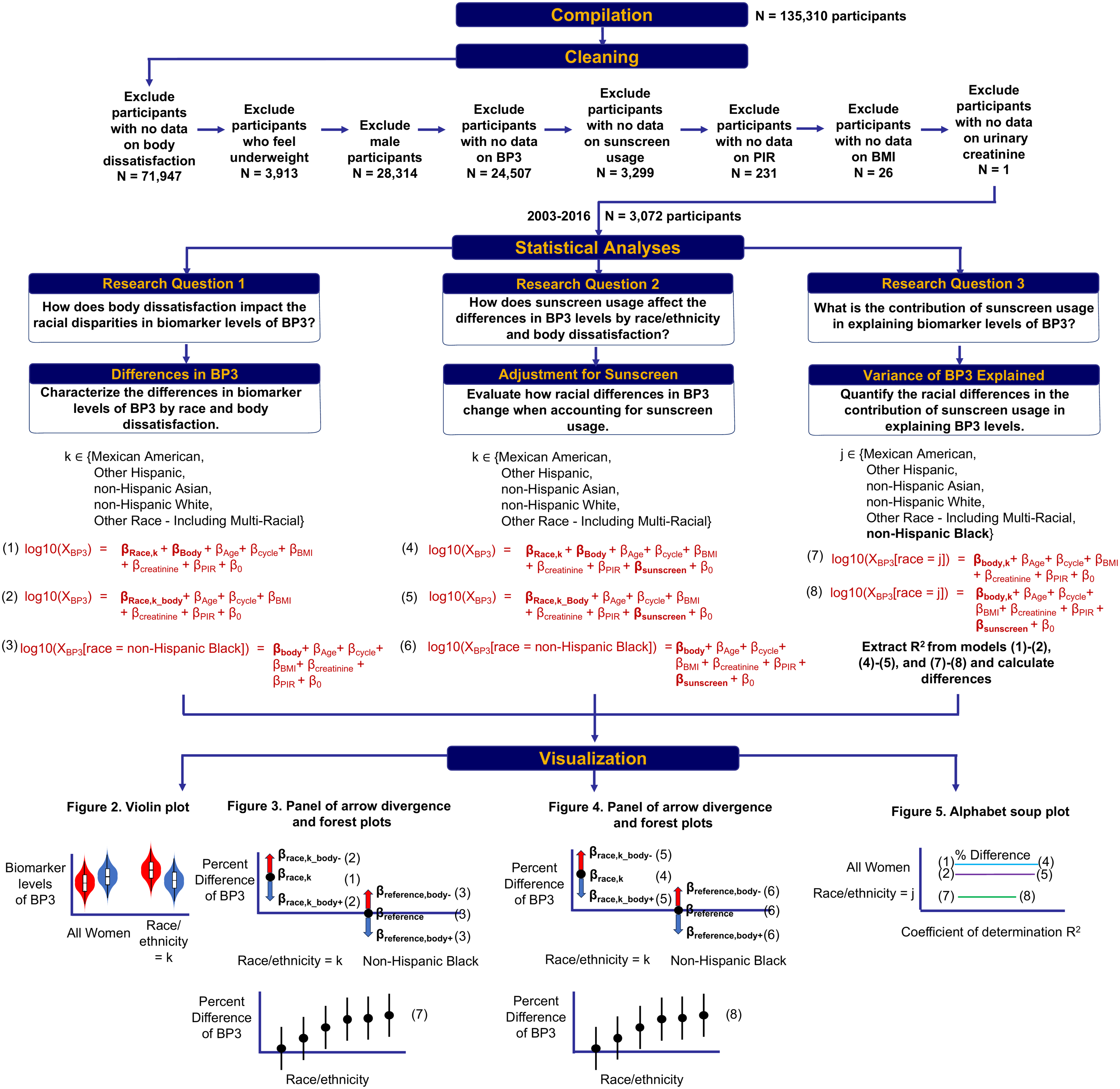

**Figure S1**. Schematic description of workflow on inclusion criteria of participants and of the statistical methods used to characterize the impact of body dissatisfaction on the racial disparities in biomarker levels of BP3. Models (1)-(2) and (4)-(5) are conducted on the NHANES women population. Models (3) and (6) are conducted specifically in non-Hispanic Black women, while Models (7) and (8) are conducted for each race.

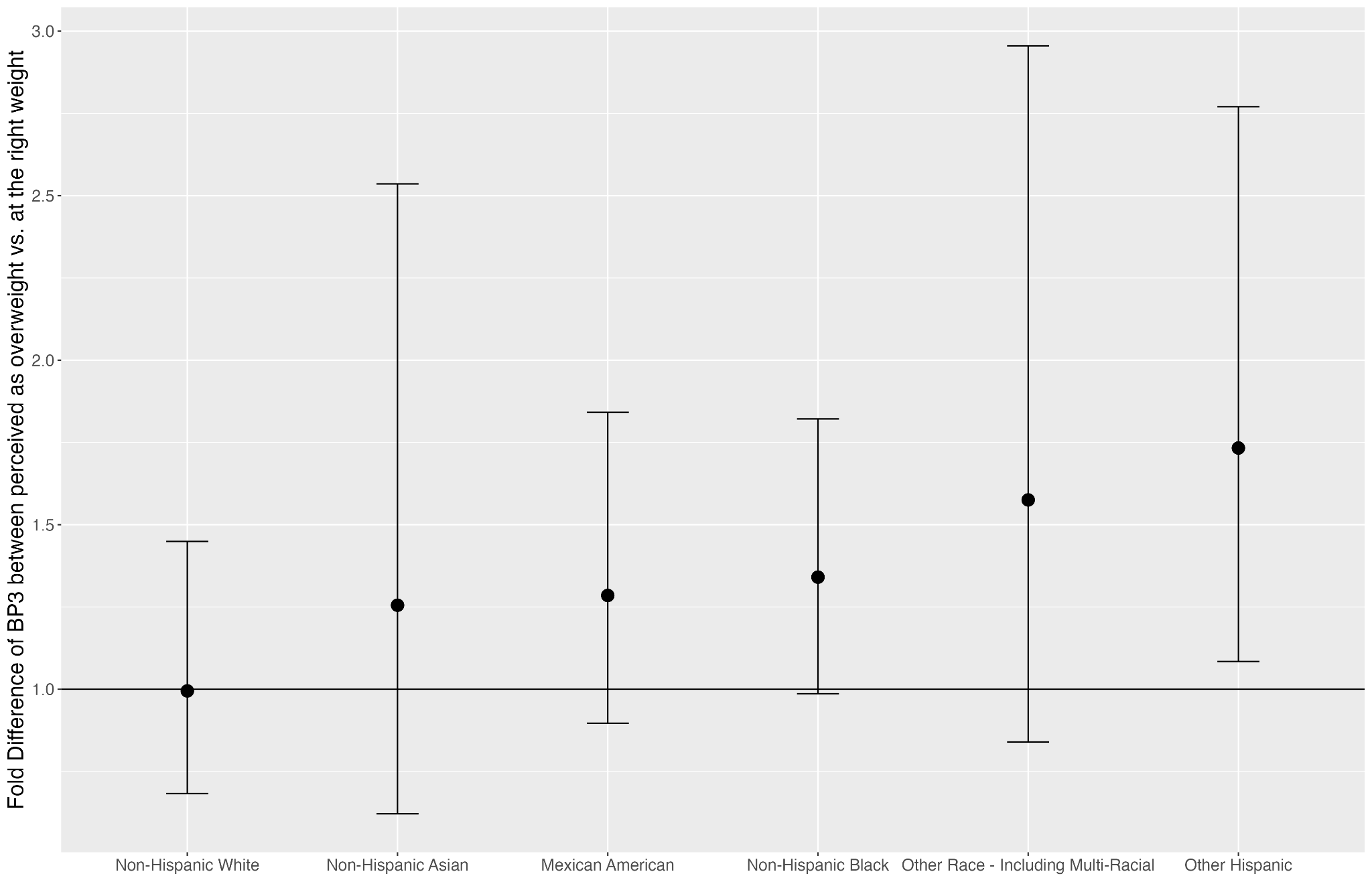

**Figure S2**. Forest plot of fold difference of BP3 biomarker levels between women who perceived themselves as overweight vs. those who perceived themselves at the right weight, **additionally adjusted for sunscreen usage**. The results are from the stratified analyses by race/ethnicity. The reference group is women who perceived themselves at the right weight in a given race/ethnicity. The results are adjusted for age, NHANES cycle, body mass index (BMI), poverty income ratio (PIR), and sunscreen usage.
